# VITAMIN D SUPPLEMENTATION FOR THE PREVENTION OF RESPIRATORY INFECTIONS IN CHILDHOOD: A SYSTEMATIC REVIEW AND META-ANALYSIS

**DOI:** 10.64898/2026.10.11.26365254

**Authors:** Hércules Kanaan Pereira Sousa, Thiago de Sousa Siqueira, Maria Adriely Machado de Castro Oliveira, Eduarda Rodrigues Belfort de Carvalho, Ana Júlia Hortegal Cavalcante, Juliana Borges Tavares da Silva, Paloma Layonara Vieira Viana, Gabriel Segrini Fidencio, Victoria Cristina Souto Costa, Lana Maria Lima Bandeira Arnaud, Alexandre Patrício Vasconcelos Matos, José Mauricio Raulino Barbosa

**Affiliations:** Centro Universitário UNINOVAFAPI (Afya Teresina), Teresina, Piauí, Brazil

**Keywords:** Vitamin D, respiratory tract infections, children, pneumonia, hospitalization, meta-analysis

## Abstract

**Background:** Vitamin D modulates innate and adaptive immunity and has been investigated for preventing pediatric respiratory infections.

**Objective:** To assess the effects of vitamin D supplementation on respiratory tract infections (RTIs), including upper RTIs, lower RTIs/recurrent pneumonia, and hospitalization/clinical severity in children and adolescents.

**Methods:** Systematic review and random-effects meta-analysis of 16 publications, of which 13 contributed to quantitative synthesis. Twenty-four effect estimates were pooled, eight per outcome. The odds ratio (OR) was analyzed on its natural logarithmic scale (logOR), using restricted maximum likelihood (REML) estimation and Knapp-Hartung (KH) adjusted inference. Where necessary, risk ratios (RRs), hazard ratios (HRs), and incidence rate ratios (IRRs) were approximated on the logOR scale.

**Results:** For respiratory infection/upper RTI, pooled logOR was −0.284 (KH-adjusted 95% confidence interval [CI]: −0.678 to 0.110; OR 0.753; p = 0.132; I^2^ = 97.2%). For lower RTI/recurrent pneumonia, logOR was −0.445 (95% CI: −0.987 to 0.097; OR 0.641; p = 0.093; I^2^ = 89.5%). For hospitalization/clinical severity, logOR was −0.609 (95% CI: −1.134 to −0.083; OR 0.544; p = 0.029; I^2^ = 78.3%).

**Conclusions:** Pooled estimates favored supplementation in direction for all three groups, although confidence intervals included the null for the first two. Heterogeneity was substantial, and the results are limited by approximations among effect measures, clinically diverse endpoints, and potentially correlated effects from individual studies.

## 1 INTRODUCTION

Vitamin D has functions that extend beyond regulation of bone and mineral metabolism, including roles in innate and adaptive immunity. Vitamin D receptors have been identified in several immune-cell populations, and their activation modulates the expression of antimicrobial peptides, including cathelicidins, which participate in host defense against respiratory pathogens. These biological mechanisms support the hypothesis that adequate vitamin D availability may influence susceptibility to respiratory infections during childhood (White, 2022; Schoenmakers, Fraser, and Forbes, 2023).

RTIs represent an important source of pediatric morbidity, particularly in the first years of life, when the immune system is still maturing and exposure to viral and bacterial agents is frequent. Recurrent episodes may lead to outpatient visits, antibiotic use, absence from school, and, in more severe cases, hospital admission or intensive care. Affordable and easily administered preventive interventions that could potentially be implemented at scale have consequently attracted interest as complementary strategies to reduce this disease burden (Lahariya *et al*., 2025; Marusca *et al*., 2023).

Vitamin D supplementation has been studied both for prevention and as adjunctive treatment. The findings have not been uniform. Some investigations reported reduced occurrence of selected respiratory infections or more severe outcomes, whereas others found no significant benefit. Loeb *et al*. (2019) reported a reduction in respiratory infections caused by non-influenza viruses but not in laboratory-confirmed influenza. Hong *et al*. (2020) described an association between regular supplementation and lower risks of RTI and lower RTI. In contrast, Somnath *et al*. (2017) observed no reduction in length of hospital stay when vitamin D was administered as adjunctive treatment to already hospitalized children.

In addition to infection incidence, investigators have assessed clinical severity, recurrence, hospitalization, safety, and serum 25-hydroxyvitamin D [25(OH)D] concentrations. This diversity in study aims, participants, dosing schedules, and supplementation regimens contributes to clinical and methodological heterogeneity. Interventions have been delivered daily, weekly, or as a single dose; participants have ranged from infants to adolescents; and supplementation has begun either before or after the onset of disease. Responses may also depend on baseline vitamin D status, dosing frequency, age, and other clinical characteristics (Marusca *et al*., 2023; Kołodziejczyk-Nowotarska, Bokiniec, and Seliga-Siwecka, 2021; Dawodu *et al*., 2019; Rosendahl *et al*., 2018).

Distinguishing prevention from treatment is particularly important. Regular prophylactic supplementation may alter nutritional status before an infectious exposure, whereas a high dose given after the development of pneumonia or a severe respiratory infection acts under different pathophysiological circumstances. Some prevention studies reported fewer respiratory episodes, whereas therapeutic use among hospitalized children did not clearly improve the clinical course. This distinction supports separating clinical contexts and outcomes in quantitative synthesis (Hong *et al*., 2020; Morris *et al*., 2021; Somnath *et al*., 2017).

Another methodological challenge is the variety of measures reported in the primary studies. The available results include RRs, HRs, IRRs, ORs, proportions, and cumulative incidences. These measures are not intrinsically interchangeable, and pooling requires careful documentation of any conversion or approximation. The present meta-analysis used logOR as its common analytical scale within random-effects models, despite variation in the measures reported by the individual studies (Loeb *et al*., 2019; Hong *et al*., 2020; Morris *et al*., 2021).

The objective of this systematic review and meta-analysis was therefore to examine the effect of vitamin D supplementation on three principal groups of pediatric respiratory outcomes: respiratory infection or upper RTI, lower RTI or recurrent pneumonia, and hospitalization or clinical severity. The statistical synthesis used random-effects models with REML estimation of between-study variance and KH-adjusted inference to account for uncertainty in the pooled estimates.

## 2 METHODS

### 2.1 Study design, research question and eligibility

This systematic review and meta-analysis was structured according to the Preferred Reporting Items for Systematic Reviews and Meta-Analyses (PRISMA 2020) guidance (Page *et al*., 2021). The question was framed in terms of population, intervention, comparator, and outcome (PICO). The population comprised infants, children, and adolescents; the intervention was vitamin D supplementation administered directly to the child or, where relevant, to the mother during pregnancy or lactation; comparators included placebo, no supplementation, another dose, or an alternative schedule; and the outcomes were respiratory events. Quantitative synthesis was organized into three categories: respiratory infection or upper RTI; lower RTI or recurrent pneumonia; and hospitalization or respiratory disease severity.

Randomized clinical trials, secondary analyses or follow-up of trials, and prospective observational studies were considered eligible when they enabled estimation of an association between vitamin D supplementation and at least one respiratory outcome. The eligible pediatric age range extended from the neonatal period to 17 years. Studies of maternal supplementation were eligible when the outcome was measured in children and the exposure contrast could be attributed to vitamin D. Case reports, letters, editorials, narrative reviews, studies without an interpretable comparator, and interventions in which the vitamin D component could not be isolated were excluded. Reports lacking quantitative respiratory outcomes were retained only for narrative discussion.

### 2.2 Search strategy and selection

The search was conducted in PubMed/MEDLINE, Google Scholar, and ScienceDirect, covering publications from 2015 to 2026. Combinations of terms related to “vitamin D”, “supplementation”, “respiratory tract infections”, “infants”, “children”, and “randomized controlled trial” were used. Reference lists of potentially relevant studies were also examined. Full texts were retrieved, when needed, through PubMed Central, journal websites, and digital object identifiers (DOIs). All records found through these strategies were considered within a single study-selection process summarized according to PRISMA 2020.

Titles and abstracts were screened for pediatric population, vitamin D intervention, and respiratory outcomes. Potentially eligible full texts were examined to confirm an appropriate comparator and sufficient quantitative information to extract or derive an effect measure with a 95% CI. When one publication reported more than one eligible result, each estimate was assigned to the corresponding outcome group. This process yielded 24 effect estimates, with eight in each group. Because several publications contributed multiple effects, the number of estimates is not equivalent to the number of independent publications.

### 2.3 Data extraction and outcome definitions

For each study, data were extracted on author, publication year, country, design, sample size, age, supplementation regimen, dose, duration, comparator, outcome definition, and available measure of association. When numbers of events and participants were available by group, they were prioritized for direct calculation of an OR. When only an RR, HR, or IRR with a 95% CI was available, the estimate and confidence limits were used to derive a logarithmic effect under the approximation described below.

The first outcome category covered respiratory infections of any cause or primarily upper respiratory infections. The second comprised lower respiratory infections, pneumonia, and recurrent pneumonia. The third covered clinical severity, including hospitalization, need for intensive care, hospitalization for lower respiratory infection, and death attributable to a respiratory episode. Length of stay, serum 25(OH)D, bone mineralization, and other non-binary outcomes were not pooled within these models.

### 2.4 Effect-measure harmonization

The selected analytical measure was logOR, with negative values indicating fewer events in the group with greater vitamin D exposure. For 2 × 2 event tables, the OR was calculated from the odds in the intervention and comparator groups, and then transformed as logOR = ln(OR); the standard error was derived from event and non-event counts. When a study reported only an RR, HR, or IRR with a 95% CI, the natural logarithm of that published ratio was used as an approximation to logOR. The standard error (SE) was estimated as SE = [ln(upper 95% CI limit) − ln(lower 95% CI limit)] / 3.92.

Importantly, RR, HR, IRR, and OR describe different estimands and are not generally mathematically equivalent. Accordingly, such transformed estimates should be regarded as approximations to the common logOR scale rather than exact odds ratios. The approximation may be especially inaccurate when events are frequent, risk varies over time, or hazards are non-proportional. When needed, reported percentages were converted to approximate event counts using available denominators. Derived estimates were retained only when the direction of the contrast and denominator were clear. Effects were oriented such that negative logOR indicated a lower frequency of adverse events with supplementation; pooled values were exponentiated for complementary presentation as ORs.

### 2.4 Statistical analysis

Three separate random-effects meta-analyses were conducted, reflecting anticipated differences in population characteristics, vitamin D dose, duration, baseline status, diagnostic definitions, and clinical setting. Between-study variance (τ^2^) was estimated using REML, with inverse-total-variance weighting. Inference about pooled effects used the KH correction (Hartung and Knapp, 2001). Results were presented as pooled logOR, corresponding exp(logOR), 95% CI, and adjusted p value; heterogeneity was summarized by τ^2^, Cochran’s Q, and I^2^.

Some publications contributed more than one estimate representing different respiratory subcategories or outcomes. As within-publication covariance matrices were unavailable for these estimates, effects from the same publication were treated as separate units in the primary models even where correlations might be present. This dependence is a methodological limitation. Forest plots are displayed on the logOR scale, with a null value of zero, in Appendices A-C. The PRISMA flow diagram is presented in Appendix D.

## 3 RESULTS

### 3.1 Study selection and quantitative synthesis

The search identified 1,720 records: 1,683 from databases and 37 from registers or other sources. Before screening, 354 duplicates and 96 records labeled as ineligible by automated tools were removed, leaving 1,270 records for title and abstract screening. A total of 1,009 records were excluded and 261 reports were sought for retrieval; seven could not be retrieved. Of 254 full-text reports assessed, 238 were excluded: ineligible population (n = 54), non-isolable vitamin D intervention (n = 41), absence of an eligible respiratory outcome (n = 46), ineligible study design (n = 33), insufficient quantitative data (n = 27), incompatible pediatric age range (n = 14), chronic disease or immunodeficiency affecting vitamin D metabolism (n = 9), extreme prematurity outside the stated criteria (n = 5), and a non-original publication, review, editorial, letter, protocol, or case report (n = 9). Sixteen publications remained in the review. The study-selection process is summarized in Figure D1 (Appendix D).

Thirteen of the 16 publications contributed directly to at least one meta-analysis. Kołodziejczyk-Nowotarska *et al*. (2021), Brustad *et al*. (2020), and Dawodu *et al*. (2019) were retained as complementary narrative evidence on safety/monitoring, bone mineralization, and adequacy of 25(OH)D, respectively. The quantitative synthesis contained 24 effect estimates: eight for respiratory infection/upper RTI, eight for lower RTI/recurrent pneumonia, and eight for hospitalization/clinical severity. Because some studies reported distinct diagnostic definitions, follow-up recurrence, or hospitalization as a component of severity, these 24 estimates do not represent 24 independent studies.

Negative logOR values indicate a lower frequency of the outcome in the group with greater vitamin D exposure, whereas zero denotes the null. All three outcome groups were synthesized using random-effects models with REML estimation and KH-adjusted inference. Table 1 summarizes the composition and Table 2 reports pooled effects.

**Table 1.** Composition of the quantitative synthesis by outcome.

| Outcome | Effect estimates (k) | Distinct publications | Studies or estimates represented |
| --- | --- | --- | --- |
| Respiratory infection / upper RTI | 8 | 8 | Loeb; Hong; Aglipay; Morris; Grant; Zhou; Ganmaa; Rosendahl |
| Lower RTI / recurrent pneumonia | 8 | 5 | Hong; Morris (3); Gupta; Chowdhury (2); Singh |
| Hospitalization / severity | 8 | 7 | Hong; Morris (2); Somnath; Singh; Labib; Chowdhury; Ganmaa |

Note: k denotes the number of effect estimates; one publication can contribute more than one estimate.

**Table 2.** Random-effects meta-analysis results (REML; KH-adjusted inference).

| Outcome | k | Pooled logOR (KH 95% CI) | OR (95% CI) | p / heterogeneity |
| --- | --- | --- | --- | --- |
| Respiratory infection / upper RTI | 8 | -0.284 (-0.678 to 0.110) | 0.753 (0.507 to 1.117) | p = 0.132; $\tau^2$ = 0.130; $I^2$ = 97.2%; Q = 247.90 |
| Lower RTI / recurrent pneumonia | 8 | -0.445 (-0.987 to 0.097) | 0.641 (0.373 to 1.102) | p = 0.093; $\tau^2$ = 0.379; $I^2$ = 89.5%; Q = 66.83 |
| Hospitalization / severity | 8 | -0.609 (-1.134 to -0.083) | 0.544 (0.322 to 0.921) | p = 0.029; $\tau^2$ = 0.299; $I^2$ = 78.3%; Q = 32.31 |

Note: Negative logOR values favor the group with greater vitamin D exposure.

### 3.2 Respiratory infection and upper respiratory tract infection

The first analysis incorporated eight estimates from studies using different supplementation schedules and definitions of respiratory infection: Loeb *et al*. (2019), Hong *et al*. (2020), Aglipay *et al*. (2017), Morris *et al*. (2021), Grant *et al*. (2015), Zhou *et al*. (2018), Ganmaa *et al*. (2020), and Rosendahl *et al*. (2018). Estimates ranged from near-null values to comparatively large reductions on the logOR scale. Hong *et al*. reported logOR −0.777 for any RTI; Zhou *et al*. reported −0.848 for influenza A; and Grant *et al*. had the most negative estimate (−2.535), with a comparatively wide interval. By contrast, Aglipay, Morris, Ganmaa, and Rosendahl reported estimates centered near zero.

The pooled random-effects logOR was −0.284 (KH-adjusted 95% CI: −0.678 to 0.110), corresponding to OR 0.753 (95% CI: 0.507 to 1.117); the adjusted p value was 0.132. Between-estimate variance was τ^2^ = 0.130, and heterogeneity was substantial (I^2^ = 97.2%; Q = 247.90). Consequently, the pooled confidence interval encompassed the null in the presence of considerable dispersion among effects. The individual estimates and pooled result are plotted in Figure A1, Appendix A.

### 3.3 Lower respiratory tract infection and recurrent pneumonia

The second analysis comprised eight estimates related to lower RTI, pneumonia, or recurrence, originating from Hong *et al*. (2020), Morris *et al*. (2021), Gupta *et al*. (2016), Chowdhury *et al*. (2021), and Singh *et al*. (2019). Morris contributed separate estimates for clinical lower RTI, microbiologically confirmed lower RTI, and microbiologically confirmed hospitalized lower RTI. Chowdhury contributed separate outcomes for any recurrent pneumonia and recurrent severe pneumonia. Thus, the number of estimates exceeded the number of independent publications.

Hong *et al*. provided the most negative estimate, logOR −1.772, for lower RTI; Singh *et al*. reported −0.687. Estimates from Morris ranged from −0.400 to −0.139, while Gupta reported 0.122 for recurrence. Chowdhury’s two effects were small or mildly negative. The pooled REML logOR was −0.445 (KH-adjusted 95% CI: −0.987 to 0.097), corresponding to OR 0.641 (95% CI: 0.373 to 1.102), p = 0.093. Between-estimate variance was τ^2^ = 0.379, with I^2^ = 89.5% and Q = 66.83. Figure B1 in Appendix B presents all eight effects.

### 3.4 Hospitalization and clinical severity

The third analysis combined eight estimates for outcomes related to hospitalization, severe lower RTI, intensive-care use, rehospitalization, and mortality. The included publications were Hong *et al*. (2020), Morris *et al*. (2021), Somnath *et al*. (2017), Singh *et al*. (2019), Labib *et al*. (2021), Chowdhury *et al*. (2021), and Ganmaa *et al*. (2020). These severity outcomes were related but were not clinically identical.

Hong *et al*. reported logOR −1.715 for RTI-related hospitalization, while Chowdhury *et al*. reported −1.620 for mortality, accompanied by a wide confidence interval because of sparse events. Morris reported −0.416 and −0.198 for two hospitalized lower RTI definitions, and estimates for Somnath, Singh, Labib, and Ganmaa ranged from −0.464 to −0.151. The pooled effect was logOR −0.609 (KH-adjusted 95% CI: −1.134 to −0.083), corresponding to OR 0.544 (95% CI: 0.322 to 0.921), p = 0.029.

Heterogeneity in this group was somewhat lower than in the other two groups but remained substantial: τ^2^ = 0.299, I^2^ = 78.3%, and Q = 32.31. The pooled interval excluded the null on the logOR scale, although the combination of heterogeneous severity outcomes and repeated estimates warrants careful interpretation. Figure C1 in Appendix C displays the contributing estimates.

## 4 DISCUSSION

This meta-analysis combined 24 effect estimates equally distributed across respiratory infection/upper RTI, lower RTI/recurrent pneumonia, and hospitalization/clinical severity. The random-effects models used REML for between-study variance and KH inference. Considerable heterogeneity was observed, reflecting differences among studies in participant age, baseline vitamin D status, dose, treatment duration, timing of supplementation, respiratory outcome definition, and clinical context (Marusca *et al*., 2023).

For general respiratory infection or upper RTI, eight estimates yielded a pooled logOR of −0.284, equivalent to an OR of approximately 0.75. Although the point estimate favored a lower event frequency with vitamin D supplementation, the KH-adjusted confidence interval included the null, and the pooled effect did not achieve conventional statistical significance. Heterogeneity was particularly high (I^2^ = 97.2%), consistent with differences in respiratory endpoints. Loeb *et al*. (2019), for instance, examined laboratory-confirmed viral infections and reported pathogen-specific effects; Hong *et al*. (2020) examined RTI occurrence in infants according to supplementation regularity. Aglipay *et al*. (2017), Ganmaa *et al*. (2020), and Rosendahl *et al*. (2018) contributed estimates close to the null.

The second group, lower RTI and recurrent pneumonia, also favored supplementation in direction (pooled logOR −0.445; OR approximately 0.64), but the interval crossed the null (p = 0.093). Heterogeneity was substantial (I^2^ = 89.5%). Hong *et al*. (2020) contributed a comparatively large negative estimate for lower RTI, while studies of pneumonia recurrence and post-hospital follow-up showed smaller or near-null effects. These differences could reflect variation in the disease stage when intervention began, baseline risk, and the clinical definition of a recurrent event (Morris *et al*., 2021; Gupta *et al*., 2016; Chowdhury *et al*., 2021; Singh *et al*., 2019).

Hospitalization and clinical severity showed the most pronounced pooled association (logOR −0.609; OR approximately 0.54), with a KH-adjusted confidence interval entirely below zero and p = 0.029. Heterogeneity remained considerable (I^2^ = 78.3%). The group combined hospital admission for respiratory illness, intensive care, severe lower RTI, rehospitalization, and mortality. Although each represents relatively severe illness, they are not equivalent endpoints. The pooled association should be interpreted in light of this composition rather than as a direct estimate of the effect on one clinically homogeneous event (Hong *et al*., 2020; Morris *et al*., 2021; Somnath *et al*., 2017; Singh *et al*., 2019; Labib *et al*., 2021; Chowdhury *et al*., 2021; Ganmaa *et al*., 2020).

The results may also depend on when vitamin D is administered. Prophylaxis given before infection can modify nutritional status over time, whereas adjunctive therapy during hospitalization begins after the disease has developed. Somnath *et al*. (2017) reported no reduction in hospital stay after a high single dose in children with severe respiratory infection, in contrast to preventive studies that reported fewer respiratory events or visits. This contrast suggests that ongoing vitamin D availability and baseline status may be more important for susceptibility than an acute dose after infection (Hong *et al*., 2020; Grant *et al*., 2015; Zhou *et al*., 2018).

The biological hypothesis for prevention remains compatible with vitamin D’s immunomodulatory roles. Receptors on immune cells contribute to the regulation of innate and adaptive responses, including expression of antimicrobial peptides. Nevertheless, a plausible mechanism does not guarantee a uniform clinical response. Baseline 25(OH)D concentrations, dose, exposure frequency, sunlight, diet, age, infecting pathogen, and population prevalence of deficiency may all influence observed effects. Such differences may help explain estimates extending from marked reductions to almost no effect (White, 2022; Schoenmakers, Fraser, and Forbes, 2023).

The choice of a common logOR scale requires particular caution. The synthesis combined effects reported as ORs, RRs, HRs, or IRRs. The logarithm of an RR, HR, or IRR cannot in general be treated as an exact logOR because each effect measure refers to a different underlying estimand. This approximation may be less problematic for rare events under restrictive conditions but introduces additional uncertainty when outcomes are common. The pooled values should therefore be interpreted as harmonized, approximated estimates, rather than as though all studies provided directly observed 2 × 2 odds ratios. A sensitivity analysis restricted to directly reported ORs or correctly convertible event data would help assess robustness.

REML and the KH adjustment were used to account for between-study variation and uncertainty in pooled effects. The KH method can be especially relevant when relatively few estimates are available. In the first two outcome groups, the pooled point estimates favored supplementation but the adjusted confidence intervals included the null. Magnitude, precision, heterogeneity, and coherence across the studies should therefore be weighed together rather than interpreting results only by p values (Hartung and Knapp, 2001).

A further challenge is the use of multiple effects from a single publication. This made it possible to represent distinct clinical and microbiological definitions, recurrent episodes, and hospitalization outcomes. However, estimates arising from overlapping samples may be correlated. Because within-publication covariance matrices were unavailable, the models treated those estimates as separate units. As a result, the eight estimates in a given outcome group do not always represent eight independent studies; confidence intervals may not fully account for this statistical dependence. Analyses selecting one effect per publication or using robust variance estimation would be informative.

Additional limitations include substantial statistical heterogeneity, broad pediatric age ranges, varied supplementation doses and schedules, pooling of preventive and therapeutic settings, and differences in baseline vitamin D status. The latter was not consistently available for stratification. Definitions of upper RTI, lower RTI, recurrent pneumonia, and clinical severity also could not be fully harmonized (Marusca *et al*., 2023).

In summary, the pooled estimates for general respiratory infections and lower RTI/recurrent pneumonia pointed toward benefit but remained compatible with no effect, whereas the pooled hospitalization/severity estimate was below the null after KH adjustment. The substantial heterogeneity, approximations among effect measures, and potential dependence among estimates limit causal interpretation. Sensitivity analyses using one estimate per publication, separating prophylactic from therapeutic supplementation, and accounting for effect dependence are warranted to examine the stability of the findings.

## 5 CONCLUSION

This systematic review and meta-analysis combined 24 reported effect estimates in three pediatric respiratory outcome groups. The pooled logOR was −0.284 (KH-adjusted 95% CI: −0.678 to 0.110) for respiratory infection/upper RTI and −0.445 (95% CI: −0.987 to 0.097) for lower RTI/recurrent pneumonia; both confidence intervals included the null. For hospitalization/clinical severity, the pooled logOR was −0.609 (95% CI: −1.134 to −0.083). Statistical heterogeneity was substantial in all three groups. The results suggest that the pooled association varies across participants, supplementation regimens, respiratory definitions, and preventive or therapeutic settings. Interpretation should account for approximations involving RR, HR, IRR, and OR, as well as multiple estimates contributed by individual publications.

## Data Availability

All data produced in the present work are contained in the manuscript

## DECLARATIONS

### Competing interests

The authors declare that they have no financial or personal competing interests related to this manuscript.

### Funding

No specific funding was reported for the preparation of this systematic review and meta-analysis.

### Data availability

The extracted and derived effect estimates used in the quantitative synthesis are presented in the manuscript tables and appendices.

### Author contributions (CRediT)

Hércules Kanaan Pereira Sousa: Conceptualization, Methodology, Formal Analysis, Data Curation, Investigation, Software, Visualization, Writing – Original Draft, Writing – Review & Editing, Project Administration.

Thiago de Sousa Siqueira: Methodology, Formal Analysis, Data Curation, Investigation, Validation, Visualization, Writing – Original Draft, Writing – Review & Editing.

Maria Adriely Machado de Castro Oliveira: Methodology, Data Curation, Investigation, Validation, Writing – Review & Editing.

Eduarda Rodrigues Belfort de Carvalho: Data Curation, Investigation, Validation, Writing – Review & Editing.

Ana Júlia Hortegal Cavalcante: Data Curation, Investigation, Validation, Writing – Review & Editing.

Juliana Borges Tavares da Silva: Investigation, Data Curation, Validation, Writing – Review & Editing.

Paloma Layonara Vieira Viana: Investigation, Data Curation, Validation, Writing – Review & Editing.

Gabriel Segrini Fidencio: Investigation, Validation, Writing – Review & Editing.

Victoria Cristina Souto Costa: Investigation, Validation, Writing – Review & Editing.

Lana Maria Lima Bandeira Arnaud: Validation, Writing – Review & Editing.

Alexandre Patrício Vasconcelos Matos: Validation, Writing – Review & Editing.

José Mauricio Raulino Barbosa: Conceptualization, Methodology, Supervision, Project Administration, Validation, Writing – Review & Editing.

### Use of generative artificial intelligence

A generative artificial intelligence tool (ChatGPT, OpenAI) was used to support editorial organization and language editing of the manuscript. Methodological decisions, source selection, data verification, and responsibility for the final content remain with the authors.

Preprint author consent: All authors have approved the current version of the manuscript and consent to its posting on medRxiv.

## APPENDIX A FOREST PLOT: RESPIRATORY INFECTION / UPPER RESPIRATORY TRACT INFECTION

**Figure A1.**
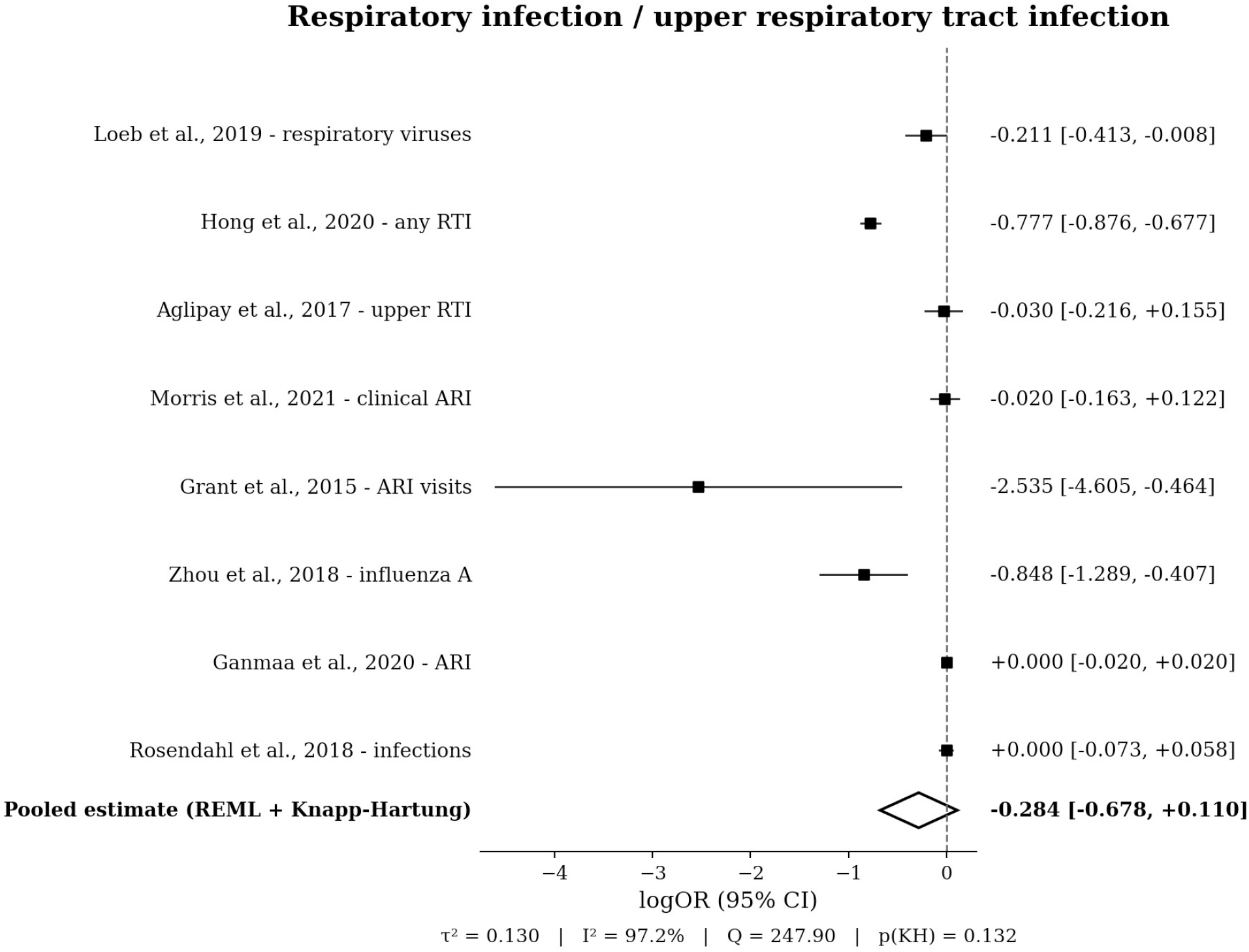
Forest plot of respiratory infection / upper respiratory tract infection. Note: Effect measure: logOR; random-effects REML model with KH-adjusted pooled inference.

## APPENDIX B FOREST PLOT: LOWER RESPIRATORY TRACT INFECTION / RECURRENT PNEUMONIA

**Figure B1.**
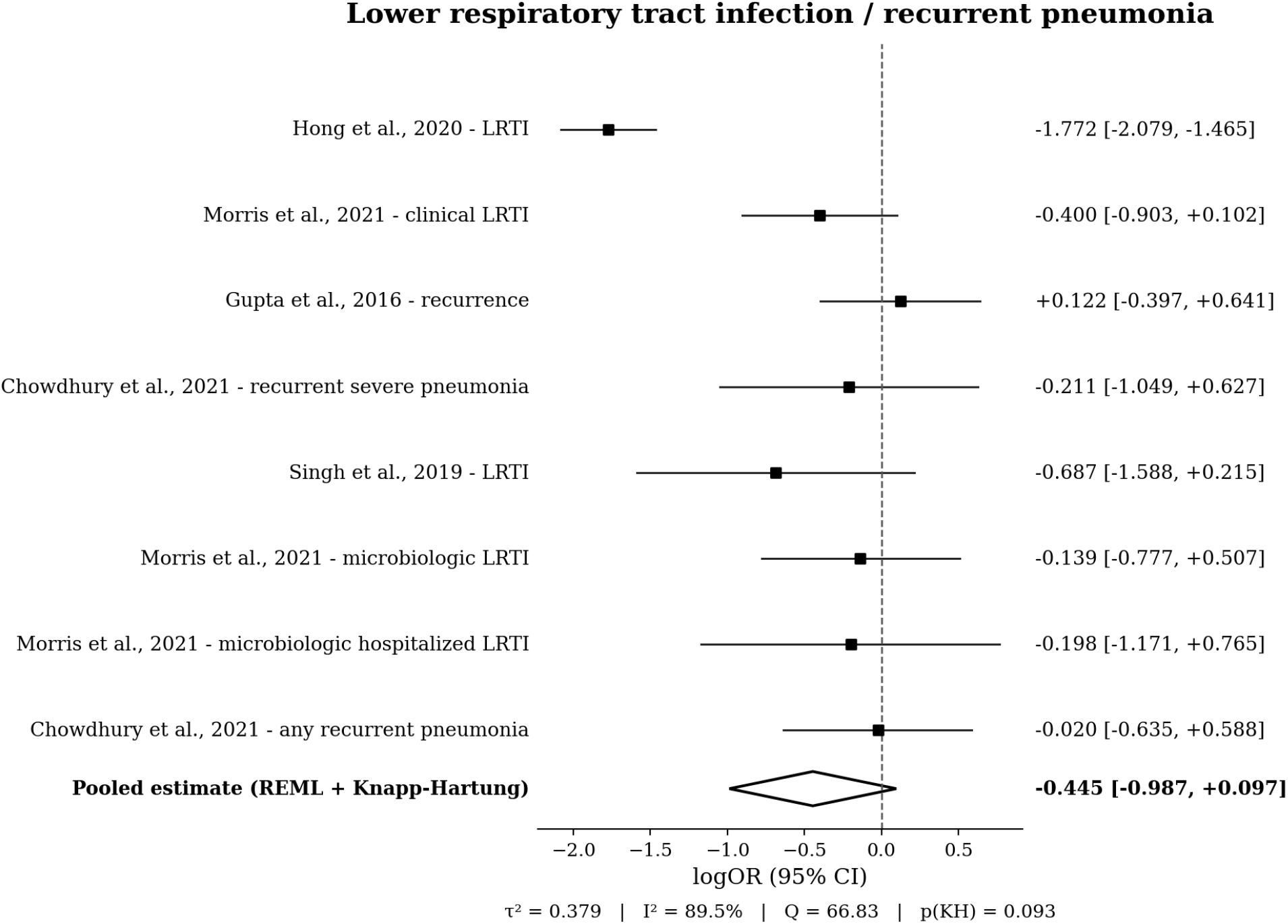
Forest plot of lower respiratory tract infection / recurrent pneumonia. Note: Effect measure: logOR; random-effects REML model with KH-adjusted pooled inference.

## APPENDIX C FOREST PLOT: HOSPITALIZATION / CLINICAL SEVERITY

**Figure C1.**
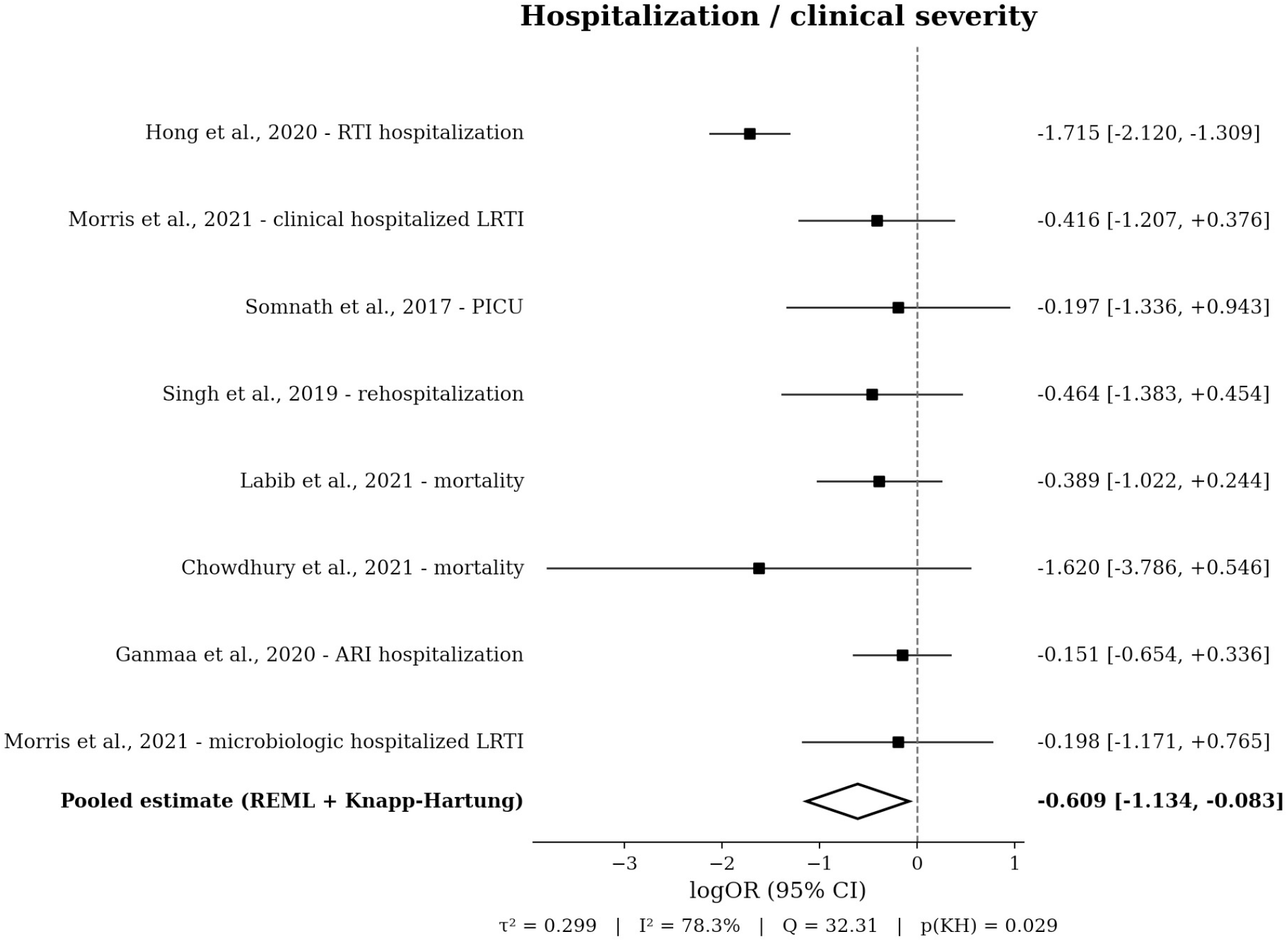
Forest plot of hospitalization / clinical severity. Note: Effect measure: logOR; random-effects REML model with KH-adjusted pooled inference.

## APPENDIX D PRISMA FLOW DIAGRAM

**Figure D1.**
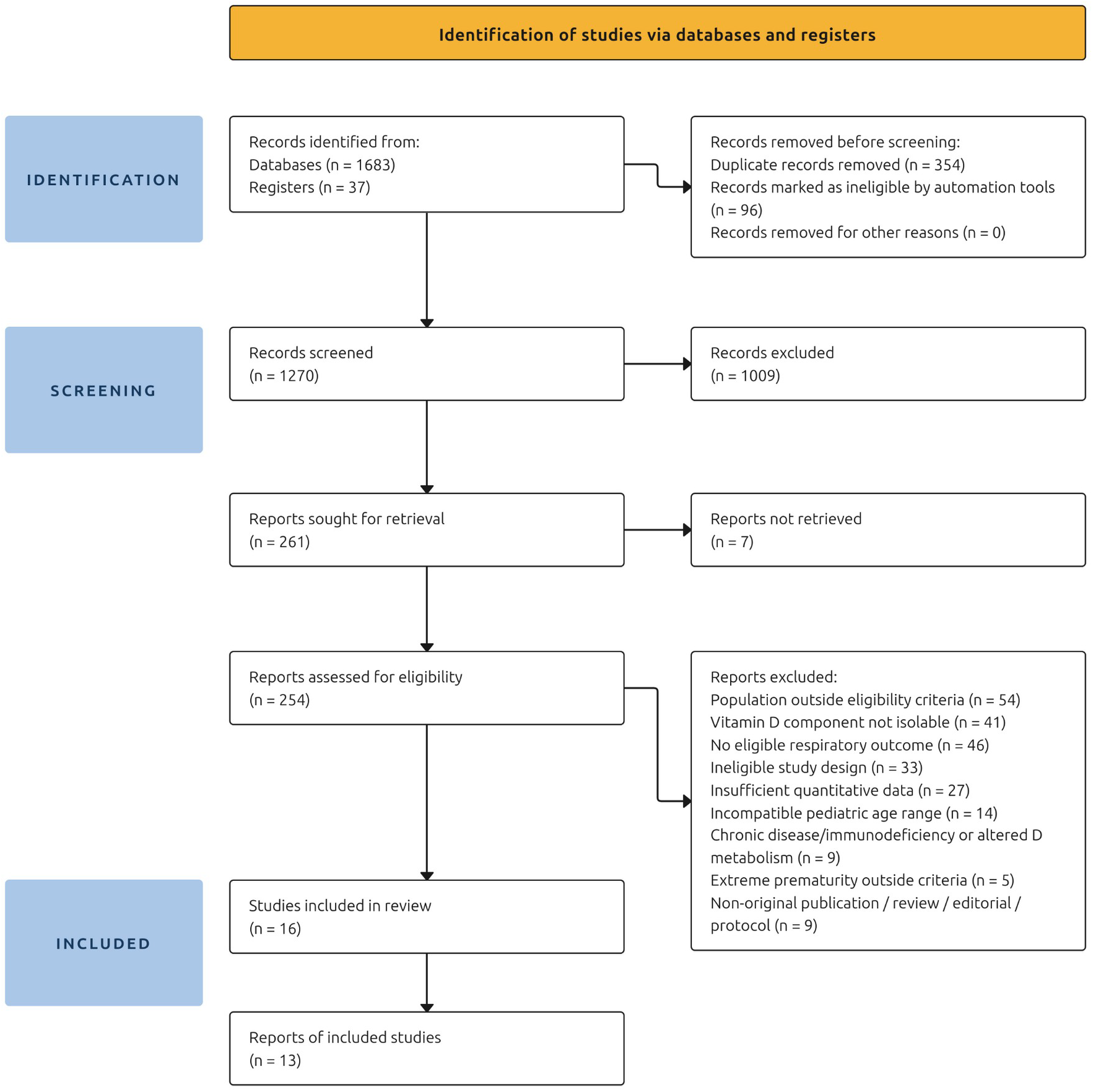
PRISMA 2020 flow diagram of study identification, screening, eligibility, inclusion in the review, and reports of included studies.

## Notes

### Competing Interest Statement

The authors have declared no competing interest.

